# Documented transboundary transmission of mpox between the Central African Republic and the Democratic Republic of the Congo

**DOI:** 10.1101/2024.08.13.24311555

**Authors:** Emmanuel Hasivirwe Vakaniaki, Eddy Kinganda-Lusamaki, Sydney Merritt, Francois Kasongo, Emile Malembi, Lygie Lunyanga, Sylvie Linsuke, Megan Halbrook, Ernest Kalthan, Elisabeth Pukuta, Adrienne Amuri Aziza, Jean Claude Makangara Cigolo, Raphael Lumembe, Gabriel Kabamba, Yvon Anta, Pierrot Bolunza, Innocent Kanda, Raoul Nganzobo, Thierry Kalonji, Justus Nsio, Patricia Matoka, Dieudonné Mwamba, Christian Ngandu, Souradet Y. Shaw, Robert Shongo, Joule Madinga, Yap Boum, Laurens Liesenborghs, Eric Delaporte, Ahidjo Ayouba, Nicola Low, Steve Ahuka Mundeke, Lisa E. Hensley, Jean-Jacques Muyembe Tamfum, Emmanuel Nakoune, Martine Peeters, Nicole A. Hoff, Jason Kindrachuk, Anne W. Rimoin, Placide Mbala-Kingebeni

## Abstract

Four confirmed mpox cases in South Ubangi province, Democratic Republic of the Congo, were linked to documented transboundary transmission from Central African Republic. Viral genome sequencing shows that the MPXV sequences belong to subclade Ia. This demonstrates the borderless nature of mpox and highlights the need for vigilant regional surveillance.

## INTRODUCTION

Mpox is a zoonotic viral infectious disease first identified in humans in the Democratic Republic of the Congo (DRC) in 1970 (1). Monkeypox virus (MPXV) is endemic in forested regions of Central and West Africa (2), subclassified into two clades: Clade I and Clade II, where subclade IIb was responsible for a global epidemic in 2022 (3). We have recommended subdivision of Clade I into subclades Ia and Ib, the latter associated with sustained human-to-human transmission in DRC (4). Clade I is endemic in Central Africa, particularly in the DRC and infections are associated with greater disease severity than Clade II (5, 6). Zoonotic spillover has been the primary driver of Clade I MPXV infections; however, sustained human-to-human transmission is increasing and spread of Clade I MPXV through sexual contact has been reported in multiple regions of the DRC (4, 7).

The DRC has faced the greatest mpox burden among endemic regions. The number of reported cases has been increasing since the cessation of smallpox vaccination campaigns in 1980 (8). This increase has accelerated from 2022 onwards (9), including into regions with no previously reported cases (10). While cross-border travel has been demonstrated as a factor involved in transmission of Clade II MPXV, less is known for Clade I cases in regional settings. Given the recent geographic expansion for Clade I MPXV and increasing observation of sustained human-to-human transmission, there are considerable concerns about geographic expansion of mpox through cross-border transmission (10).

The Central African Republic (CAR) shares a >1,000-mile border with DRC. CAR has reported 40 confirmed mpox outbreaks (95 suspected from 2001-2021), increasing from 0-2 annually from 2001-2017 to nine annually since 2018 (11). Most of these outbreaks occurred in two regions along the DRC border. Here, we report suspected cross-border transmission of mpox between CAR and DRC in 2023 and potential links with MPXV circulation in DRC.

## THE STUDY

An alert for a suspected mpox case was issued by the South Ubangi Provincial Health Division, DRC, in January 2023 following the death of a fisherman who presented with respiratory distress and skin lesions in Mbaya Health Zone. South Ubangi Province directly borders CAR, separated by the Ubangi River. The National Programme for Control of Mpox and Viral Haemorrhagic Fevers (PNLMPX-VHF) and the National Institute for Biomedical Research (INRB) deployed a multidisciplinary team to investigate the case with the South Ubangi Provincial Health Division. As part of the investigation, the national and provincial health teams conducted supplemental training sessions to raise awareness of the clinical signs and transmission modes for mpox.

**Figure 1** shows a documented chain of MPXV transmission from CAR to Mbaya Health Zone. The index case was an adult old male resident of Bangui, CAR, who moved frequently between CAR and DRC via the Ubangi River (cluster 1, case 1: C1). Following suspected exposure to wildlife meat and the potential consumption of rodents in CAR in early January 2023, with symptom onset began shortly after (day 0) and including fever, headache, chills, and cutaneous rash. After traveling to the DRC, he was admitted to Mbaya General Reference Hospital two days later, where he died on day 9 post-symptom onset after experiencing respiratory distress. No samples were collected from this probable mpox case nor any information on his immune status.

**Figure 1.**
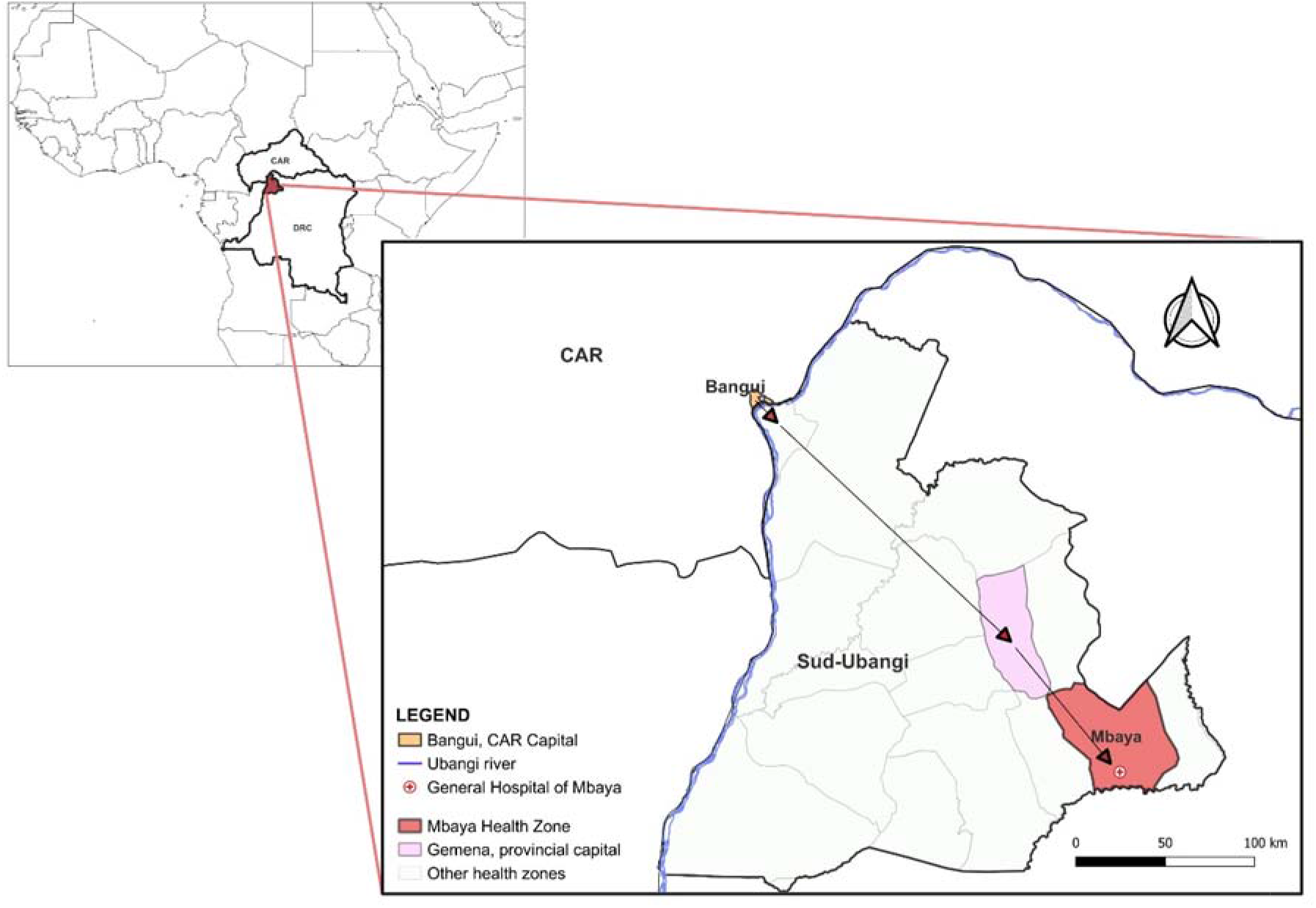
Suggested transmission route of MPXV between Bangui, Central African Republi (CAR) and Mbaya Health Zone, South Ubangi Province, Democratic Republic of the Congo, January 2023

Fifteen contacts of the patient were identified in Mbaya Health Zone, including five health care workers and 10 close contacts (four close friends and six family contacts). The investigation revealed a cluster of three symptomatic family contacts. Two had samples taken, which were positive for MPXV by PCR; the adult female partner of C1 (C2) and their child (C3). A second child of C1 and C2 (C4) was treated locally for suspected mpox through traditional methods and was not investigated further. No suspected mpox cases were identified among family or close contacts of C1 in Bangui, CAR.

During the investigation, a second mpox cluster was identified, which likely originated from a different source. Three suspected mpox cases were identified at the same hospital and at the same time as the first cluster, with no known epidemiologic link. An adult female patient (cluster 2, case 1: D1) was hospitalized for nonspecific signs of mpox at the same time as C1 and reported contact with a child with mpox symptoms ∼28 days prior. The two additional suspected cases were the child of D1 (D2), with subsequent PCR-confirmed mpox, and a contact of D2 (D3) who tested PCR negative. **(Figure 2**).

**Figure 2.**
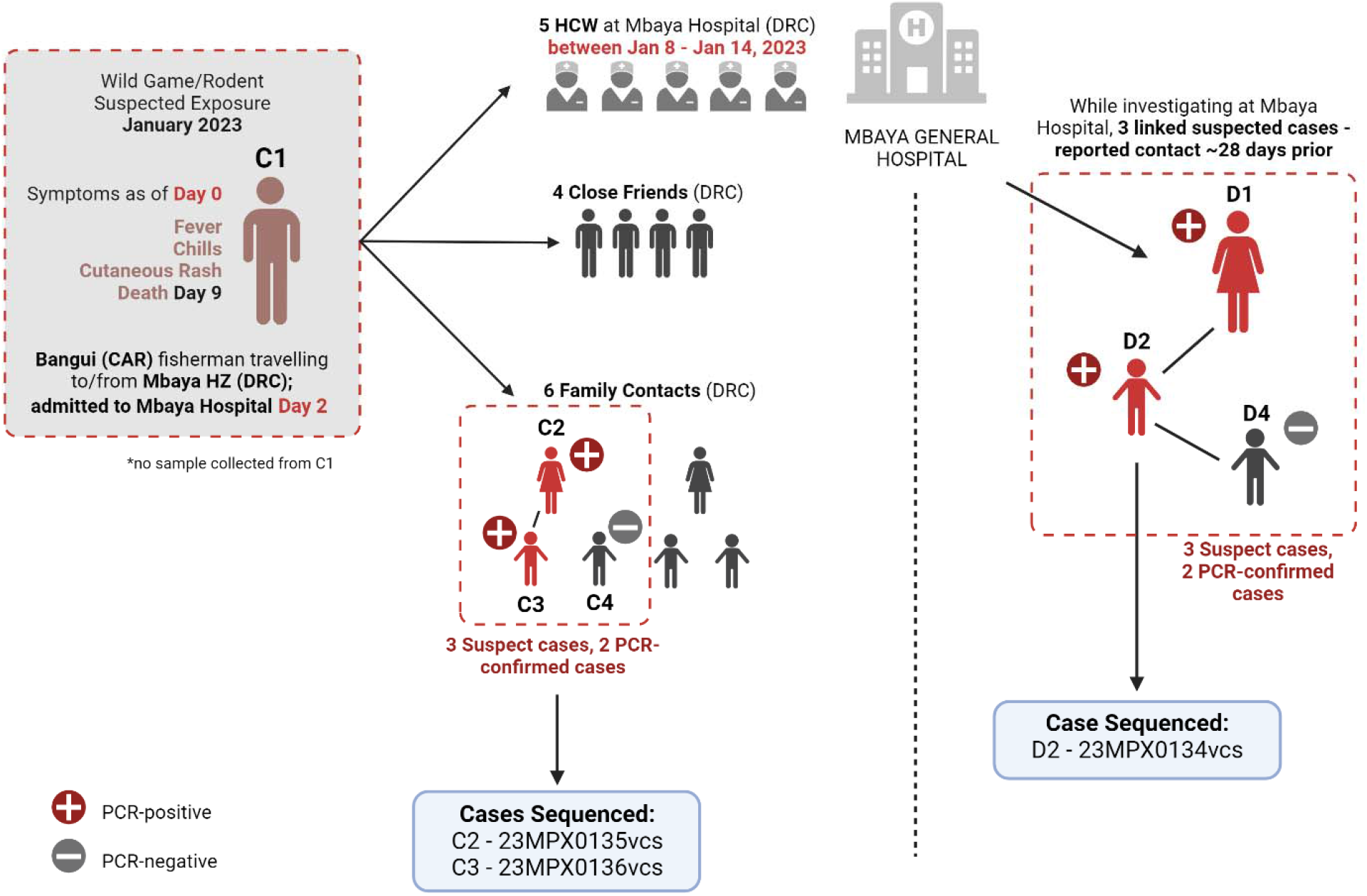
Suggested transmission chain from Central African Republic to the Democratic Republic of the Congo and second identified chain at Mbaya General Reference Hospital, DRC. Abbreviations: HCW, healthcare workers; HZ, health zone

Confirmed mpox clinical symptoms among the confirmed cases included multiple pustular and papular lesions (C2), disseminated pustules (C3), discreet lacrimation in the left eye and post-inflammatory hyperpigmentation (D1), and hyperpigmented, disseminated macules and steep-edged ulcerations (D2).

Subsequent viral genome sequencing was performed on three PCR-positive samples: two family members in the first mpox cluster (C2 and C3) and one from the second cluster (D2) **(Figure 3)**. Phylogenetic analysis included 98 clade I MPXV previously published genomes. The phylogenetic tree was generated by maximum likelihood using the K3Pu+F+I model (12, 13). The three sequences, all clade Ia, clustered with MPXV genome sequences isolated from CAR. The genetic distance between the new cases and the date between the CAR and DRC samples is too long to conclude direct links between the two countries. Nevertheless, the new samples from South-Ubangi cluster in the same subgroup (group II) according to previous classification of MPXV sequences in clade Ia (14).

**Figure 3:**
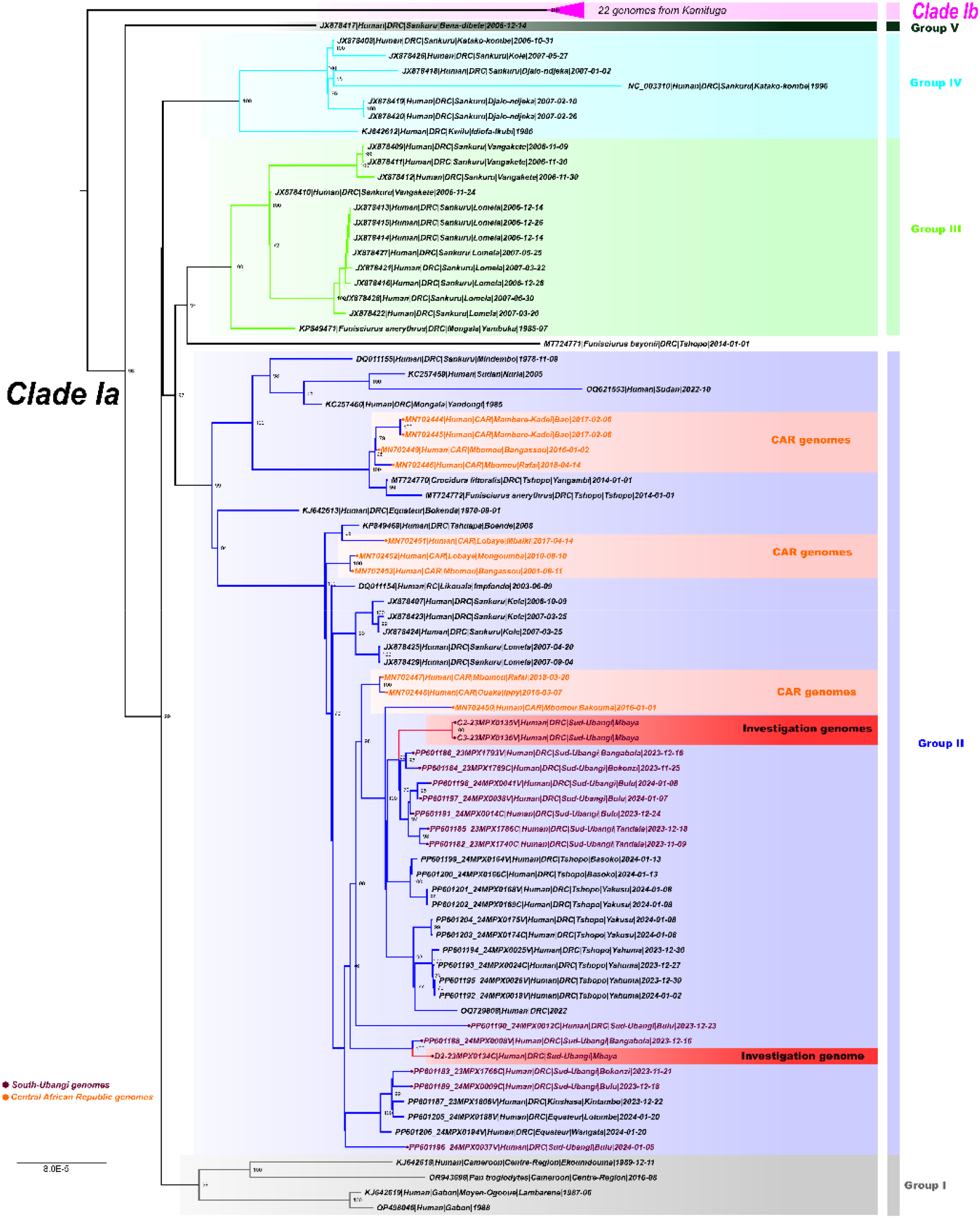
Phylogenetic analysis of MPXV sequences from Mbaya rural health zone. Phylogenetic analysis of MPXV genome sequences from samples described in this study and clade I MPXV sequences from Central Africa. Scale bar is in substitutions per site; bootstrap support values are shown at branch points. DNA was extracted at INRB using a Qiagen DNA Mini kit from blood samples and subsequently screened for MPXV with an Orthopoxvirus-specific real-time PCR assay. Whole genome sequencing was attempted on samples from the index case by Next Generation Sequencing (NGS). The library preparation was performed using Illumina DNA Prep with Enrichment and the libraries were enriched for MPXV using biotinylated custom probes synthesized by Twist Biosciences. Note that 23MPX0134C(D2), 23MPX0135V(C2), and 23MPX0136V(C3) are samples from crust or vesicles from separate individuals.

The PNLMPX-VHF investigated a further 109 suspected mpox cases from 15 health zones in South Ubangi province from January to November 2023. Cases were predominantly male (60 males; 46 females; 3 unavailable) with an average age of 21 years (range 35 months to 63 years). Of these, 61 cases (56 %) were confirmed positive by PCR at INRB. Three mpox negative cases were confirmed positive for varicella zoster virus (3/48; 6%).

## DISCUSSION

This investigation describes two clusters of mpox cases in Mbaya health zone in South Ubangi province, DRC, presumably resulting from two different introductions. Epidemiological links to cross-border travel from CAR were found in the first cluster and genomic links with previous cases from CAR for both. MPXV transmission in DRC is currently driven by both zoonotic and human to human contact, thus increasing the complexity of containment and mitigation efforts. There are considerable concerns about wider regional and international expansion of mpox including the concentration of MPXV outbreaks in CAR and DRC along the Ubangi River, recent increases in sustained human-to-human transmission, resource limitations for identification and treatment of mpox, disease stigma, population displacement due to conflict, and transient cross-border transit.

In August 2022, a first regional meeting involving six neighbouring countries of Central and West Africa was held in Kinshasa, DRC to establish a regional mpox surveillance network - the Mpox Threat Reduction Network. Through this network multiple teams that included experts from the Health Ministries of the DRC and CAR which were able to communicate rapidly on suspected cases crossing borders in South Ubangi while also reporting to the International Health Regulations focal point in the DRC and CAR. This investigation highlights the importance of long-term collaborative partnerships for sustained mpox surveillance and containment in endemic regions.

## Data Availability

All data produced in the present study are available upon reasonable request to the authors

## ACKNOWLEDGEMENTS

Figures included in this manuscript were generated using Biorender.com.

## REFERENCES

1. Ladnyj ID, Ziegler P, Kima E. A human infection caused by monkeypox virus in Basankusu Territory, Democratic Republic of the Congo. Bull World Health Organ. 1972;46(5):593–7.

2. Titanji BK, Tegomoh B, Nematollahi S, Konomos M, Kulkarni PA. Monkeypox: A Contemporary Review for Healthcare Professionals. Open Forum Infect Dis. 2022 Jul;9(7):ofac310.

3. Happi C, Adetifa I, Mbala P, Njouom R, Nakoune E, Happi A, et al. Urgent need for a non-discriminatory and non-stigmatizing nomenclature for monkeypox virus. PLoS Biol. 2022 Aug;20(8):e3001769.

4. Vakaniaki EH, Kacita C, Kinganda-Lusamaki E, O’Toole A, Wawina-Bokalanga T, Mukadi-Bamuleka D, et al. Sustained Human Outbreak of a New MPXV Clade I Lineage in the Eastern Democratic Republic of the Congo. Nat Med. 2024 Jun 13.

5. Weaver JR, Isaacs SN. Monkeypox virus and insights into its immunomodulatory proteins. Immunol Rev. 2008 Oct;225:96–113.

6. McCollum AM, Damon IK. Human monkeypox. Clin Infect Dis. 2014 Jan;58(2):260–7.

7. Kibungu EM, Vakaniaki EH, Kinganda-Lusamaki E, Kalonji-Mukendi T, Pukuta E, Hoff NA, et al. Clade I-Associated Mpox Cases Associated with Sexual Contact, the Democratic Republic of the Congo. Emerg Infect Dis. 2024 Jan;30(1):172–6.

8. MSPHP. REPORT ON THE EPIDEMIOLOGICAL SITUATION OF MONKEYPOX VIRUS—S1-S6 2024. Ministry of Public Health, Hygiene and Prevention of the Democratic Republic of the Congo; 2024.

9. WHO. Disease Outbreak News: Mpox - Democratic Republic of the Congo; 2024 14 June 2024.

10. Mbala-Kingebeni P, Rimoin AW, Kacita C, Liesenborghs L, Nachega JB, Kindrachuk J. The time is now (again) for mpox containment and elimination in Democratic Republic of the Congo. PLOS Glob Public Health. 2024;4(6):e0003171.

11. Besombes C, Mbrenga F, Schaeffer L, Malaka C, Gonofio E, Landier J, et al. National Monkeypox Surveillance, Central African Republic, 2001-2021. Emerg Infect Dis. 2022 Dec;28(12):2435–45.

12. Kalyaanamoorthy S, Minh BQ, Wong TKF, von Haeseler A, Jermiin LS. ModelFinder: fast model selection for accurate phylogenetic estimates. Nat Methods. 2017 Jun;14(6):587–9.

13. Minh BQ, Schmidt HA, Chernomor O, Schrempf D, Woodhams MD, von Haeseler A, et al. IQ-TREE 2: New Models and Efficient Methods for Phylogenetic Inference in the Genomic Era. Mol Biol Evol. 2020 May 1;37(5):1530–4.

14. Berthet N, Descorps-Declere S, Besombes C, Curaudeau M, Nkili Meyong AA, Selekon B, et al. Genomic history of human monkey pox infections in the Central African Republic between 2001 and 2018. Sci Rep. 2021 Jun 22;11(1):13085.

